# Persistence of symptoms up to 10 months following acute COVID-19 illness

**DOI:** 10.1101/2021.03.07.21253072

**Authors:** Roy H. Perlis, Jon Green, Mauricio Santillana, David Lazer, Katherine Ognyanova, Matthew Simonson, Matthew A. Baum, Alexi Quintana, Hanyu Chwe, James Druckman, John Della Volpe, Jennifer Lin

## Abstract

**Importance:** COVID-19 symptoms are increasingly recognized to persist among a subset of individual following acute infection, but features associated with this persistence are not well-understood.

**Objective:** We aimed to identify individual features that predicted persistence of symptoms over at least 2 months at the time of survey completion.

Design: Non-probability internet survey. Participants were asked to identify features of acute illness as well as persistence of symptoms at time of study completion. We used logistic regression models to examine association between sociodemographic and clinical features and persistence of symptoms at or beyond 2 months.

**Setting:** Ten waves of a fifty-state survey between June 13, 2020 and January 13, 2021.

**Participants:** 6,211 individuals who reported symptomatic COVID-19 illness confirmed by positive test or clinician diagnosis.

**Exposure:** symptomatic COVID-19 illness

**Results:** Among 6,211 survey respondents reporting COVID-19 illness, with a mean age of 37.8 (SD 12.2) years and 45.1% female, 73.9% white, 10.0% Black, 9.9% Hispanic, and 3.1% Asian, a total of 4946 (79.6%) had recovered within less than 2 months, while 491 (7.9%) experienced symptoms for 2 months or more. Of the full cohort, 3.4% were symptomatic for 4 months or more and 2.2% for 6 months or more. In univariate analyses, individuals with persistent symptoms on average reported greater initial severity. In logistic regression models, older age was associated with greater risk of persistence (OR 1.10, 95% CI 1.01-1.19 for each decade beyond 40); otherwise, no significant associations with persistence were identified for gender, race/ethnicity, or income. Presence of headache was significantly associated with greater likelihood of persistence (OR 1.44, 95% CI 1.11-1.86), while fever was associated with diminished likelihood of persistence (OR 0.66, 95% CI 0.53-0.83).

**Conclusion and Relevance:** A subset of individuals experience persistent symptoms from 2 to more than 10 months after acute COVID-19 illness, particularly those who recall headache and absence of fever. In light of this prevalence, strategies for predicting and managing such sequelae are needed.

**Trial Registration:** NA

**Key Points:** *Question:* Which individuals are at greatest risk for post-acute sequelae of COVID-19?

*Findings:* In this non-probability internet survey, among 6,211 individuals with symptomatic COVID-19 illness, 7.9% experienced persistence of symptoms lasting 2 months or longer. Older age, but not other sociodemographic features, was associated with risk for persistence, as was headache.

*Meaning:* Identifying individuals at greater risk for symptomatic persistence may facilitate development of targeted interventions.

## INTRODUCTION

Postviral syndromes following epidemics have been recognized for more than a century, and COVID-19 appears to be no exception^1^. For a subset of individuals with acute Coronavirus-19 (COVID-19) disease, symptoms may persist beyond a month^2^, with some patients reporting symptoms at least 6 months later. At 60 days after hospitalization, one study found that 87% of individuals with COVID-19 had at least one persistent symptom^3^, while at 6 months post-hospitalization, a large study found 63% of participants experienced fatigue or weakness, and 26% experienced sleep disturbance^4^. The prevalence of such symptoms among less severely ill patients is less well-characterized; one app-based study including non-hospitalized individuals found rates of persistence of 4.5% at 2 months^5^.

Numerous aspects of this symptom persistence, referred to as post-acute sequelae of COVID-19, remain poorly understood, with a recent evidence review suggesting this phenomenon may actually reflect multiple different syndromes(1). In particular, it is not known which individuals will experience full recovery, and which persistence. A particular concern is whether disadvantaged groups may be disproportionately impacted, as they have been by acute infection^7^. On the other hand, if high-risk individuals could be identified, it might be possible to develop strategies to mitigate or prevent symptom persistence, prompting calls for increased emphasis on investigation of post-acute sequelae of COVID-19^8^.

In order to better characterize symptom persistence, we utilized data from a multi-wave US survey that included questions about COVID-19 encompassing 50 states and the District of Columbia. Importantly, the survey did not focus only on COVID-19, yielding less likelihood of selection bias than more focused surveys of COVID-19 persistence where participants opt in. We aimed to identify individual features that predicted persistence of symptoms over at least 2 months at the time of survey completion.

## METHOD

### Study Design

We conducted 10 waves of an online survey between June 13, 2020 and January 10, 2021 across 50 states and the District of Columbia, applying nonprobability sampling using representative quotas in order to balance age, gender, and race/ethnicity. These waves included a total of 124962 non-overlapping individuals. The study was determined to be exempt by the Institutional Review Board of Harvard University; all participants signed consent online prior to survey access.

### Measures

All respondents were asked if they had received a positive COVID-19 test result, and/or had received a diagnosis of likely COVID-19 by a clinician. Those who endorsed either of these completed a checklist to identify symptoms present during initial illness, and those who endorsed persistent symptoms at the time of survey completion completed a second checklist with an expanded list of symptoms. The full survey is available at [link]^1^.

### Analysis

In primary analysis, we examined all individuals with either a positive test or clinician diagnosis of COVID-19 at any point, consistent with an evidence review indicating a high proportion of individuals lacked access to testing earlier in the pandemic^6^; sensitivity analyses planned a priori examined a narrower definition based on a positive test alone. Individual symptoms were characterized in terms of prevalence at acute infection, and then among those with persistent symptoms. Duration of illness in months was estimated based on date of survey completion and reported month of initial illness and subsequent months reported to be symptomatic. (Individuals who remained symptomatic at time of survey completion, with symptom duration of less than 2 months, were excluded from analysis of persistence). We utilized logistic regression in R 4.0^9^ to examine association of symptoms during acute infection with risk for persistence, adjusted for age in decade, gender, race/ethnicity indicated on a 5-item questionnaire, income decile, and rural/suburban/urban area of residence, as well as self-reported severity of initial illness on a 4-point scale (‘not at all’, ‘not too’, ‘somewhat’, or ‘very’ severe).

## RESULTS

The full cohort included 6,211 individuals who reported symptomatic COVID-19 illness confirmed by positive test or clinician diagnosis (Table 1), with a mean age of 37.8 (SD 12.2) years, who were 45.1% female, 73.9% white, 10.0% Black, 9.9% Hispanic, and 3.1% Asian. Among them, a total of 4946 (79.6%) had recovered within less than 2 months, while 491 (7.9%) experienced symptoms for 2 months or more, and 774 (12.5%) were not yet recovered but had been ill for less than 2 months. Of the full cohort, 3.4% were symptomatic for 4 months or more and 2.2% for 6 months or more. Figure 1 illustrates time to recovery, right-censoring individuals still ill at month of survey.

**Table 1.** Features of individuals who did or did not report COVID-19 symptom persistence

|  | Resolution in 0-1 month (N=4946) | Persistence 2+ months (N=491) | Total (N=5437) | p value |
| --- | --- | --- | --- | --- |
| **Positive test (a)** | 3424 (69.8%) | 332 (68.3%) | 3756 (69.7%) | 0.482 |
| **Clinician diagnosis** | 4565 (92.3%) | 449 (91.4%) | 5014 (92.2%) | 0.502 |
| **Age (SD)** | 37.81 (11.88) | 38.46 (12.33) | 37.87 (11.92) | 0.246 |
| **Gender (Female)** | 2031 (41.1%) | 217 (44.2%) | 2248 (41.3%) | 0.179 |
| **Income decile (b )** | 8 | 0 | 8 | 0.03 |
| **0** | 21 (0.4%) | 3 (0.6%) | 24 (0.4%) |  |
| **1** | 470 (9.5%) | 63 (12.8%) | 533 (9.8%) |  |
| **2** | 367 (7.4%) | 49 (10.0%) | 416 (7.7%) |  |
| **3** | 350 (7.1%) | 43 (8.8%) | 393 (7.2%) |  |
| **4** | 369 (7.5%) | 36 (7.3%) | 405 (7.5%) |  |
| **5** | 632 (12.8%) | 66 (13.4%) | 698 (12.9%) |  |
| **6** | 608 (12.3%) | 44 (9.0%) | 652 (12.0%) |  |
| **7** | 847 (17.2%) | 78 (15.9%) | 925 (17.0%) |  |
| **8** | 635 (12.9%) | 59 (12.0%) | 694 (12.8%) |  |
| **9** | 639 (12.9%) | 50 (10.2%) | 689 (12.7%) |  |
| **Race** |  |  |  | 0.102 |
| **White** | 3697 (74.7%) | 346 (70.5%) | 4043 (74.4%) |  |
| **Hispanic** | 481 (9.7%) | 46 (9.4%) | 527 (9.7%) |  |
| **Black** | 470 (9.5%) | 62 (12.6%) | 532 (9.8%) |  |
| **Asian** | 153 (3.1%) | 17 (3.5%) | 170 (3.1%) |  |
| **Other** | 145 (2.9%) | 20 (4.1%) | 165 (3.0%) |  |
| **Residence** |  |  |  | 0.071 |
| **Rural** | 688 (13.9%) | 74 (15.1%) | 762 (14.0%) |  |
| **Suburban** | 2521 (51.0%) | 270 (55.0%) | 2791 (51.3%) |  |
| **Urban** | 1737 (35.1%) | 147 (29.9%) | 1884 (34.7%) |  |
| **Severity (c )** |  |  |  | < 0.001 |
| **Not at all** | 724 (14.6%) | 31 (6.3%) | 755 (13.9%) |  |
| **Not too** | 1337 (27.0%) | 84 (17.2%) | 1421 (26.2%) |  |
| **Somewhat** | 1673 (33.8%) | 190 (38.9%) | 1863 (34.3%) |  |
| **Very** | 1210 (24.5%) | 184 (37.6%) | 1394 (25.7%) |  |
| **Symptoms** |  |  |  |  |
| **Fever** | 2434 (49.2%) | 241 (49.1%) | 2675 (49.2%) | 0.957 |
| **Chills** | 1516 (30.7%) | 209 (42.6%) | 1725 (31.7%) | < 0.001 |
| **Shaking** | 1392 (28.1%) | 201 (40.9%) | 1593 (29.3%) | < 0.001 |
| **Congestion** | 2305 (46.6%) | 262 (53.4%) | 2567 (47.2%) | 0.004 |
| **Muscle Pain** | 1574 (31.8%) | 223 (45.4%) | 1797 (33.1%) | < 0.001 |
| **Cough** | 1778 (35.9%) | 199 (40.5%) | 1977 (36.4%) | 0.044 |
| **Sore Throat** | 1966 (39.7%) | 226 (46.0%) | 2192 (40.3%) | 0.007 |
| **Headache** | 1036 (20.9%) | 167 (34.0%) | 1203 (22.1%) | < 0.001 |
| **Shortness of Breath** | 1682 (34.0%) | 203 (41.3%) | 1885 (34.7%) | 0.001 |
| **Loss of Taste/Smell** | 2437 (49.3%) | 269 (54.8%) | 2706 (49.8%) | 0.02 |

a. missing (44 short, 5 long)
b. missing (8 short, 0 long)
c. missing (2 short, 2 long)

**Figure 1.**
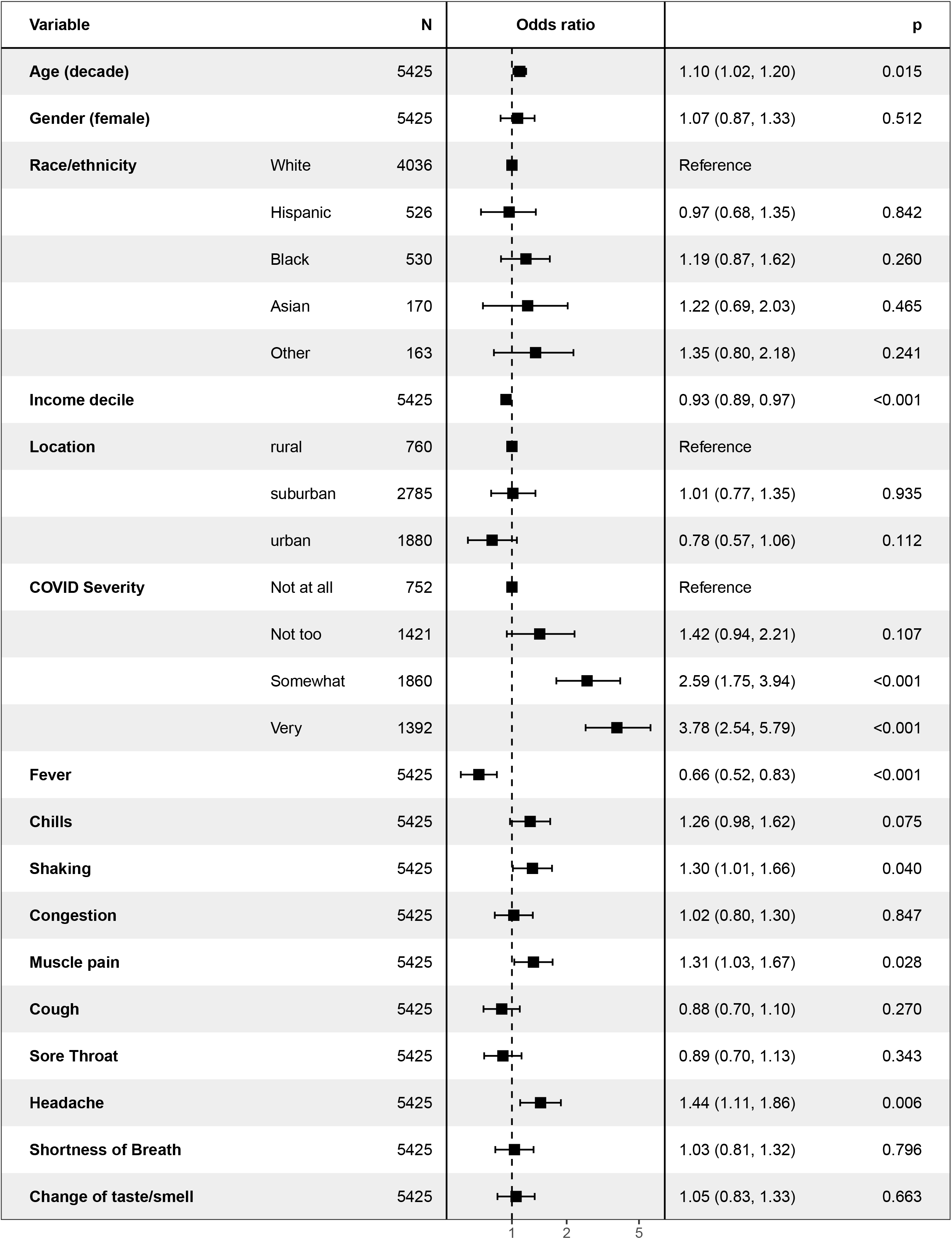
Among those with prior self-reported COVID-19 illness, logistic regression model of risk for persistence

We next examined features associated with persistent symptoms (eTable 1). In univariate analyses, individuals with persistent symptoms on average reported greater initial severity, greater likelihood of confirmation of diagnosis by a clinician, and greater prevalence of all individual symptoms except fever (Table 1).

In multiple logistic regression (Figure 1), age was associated with greater risk (for each decade beyond age 40, OR 1.10, 95% CI 1.02-1.20). Otherwise, no significant associations with persistence were identified among sociodemographic features including gender, race/ethnicity, and income decile. Among individual symptoms during acute illness, headache was significantly associated with greater likelihood of persistence (Figure 2), while fever was associated with diminished likelihood of persistence. In addition, greater overall self-reported severity during acute illness was also associated with likelihood of persistence.

**Figure 2.**
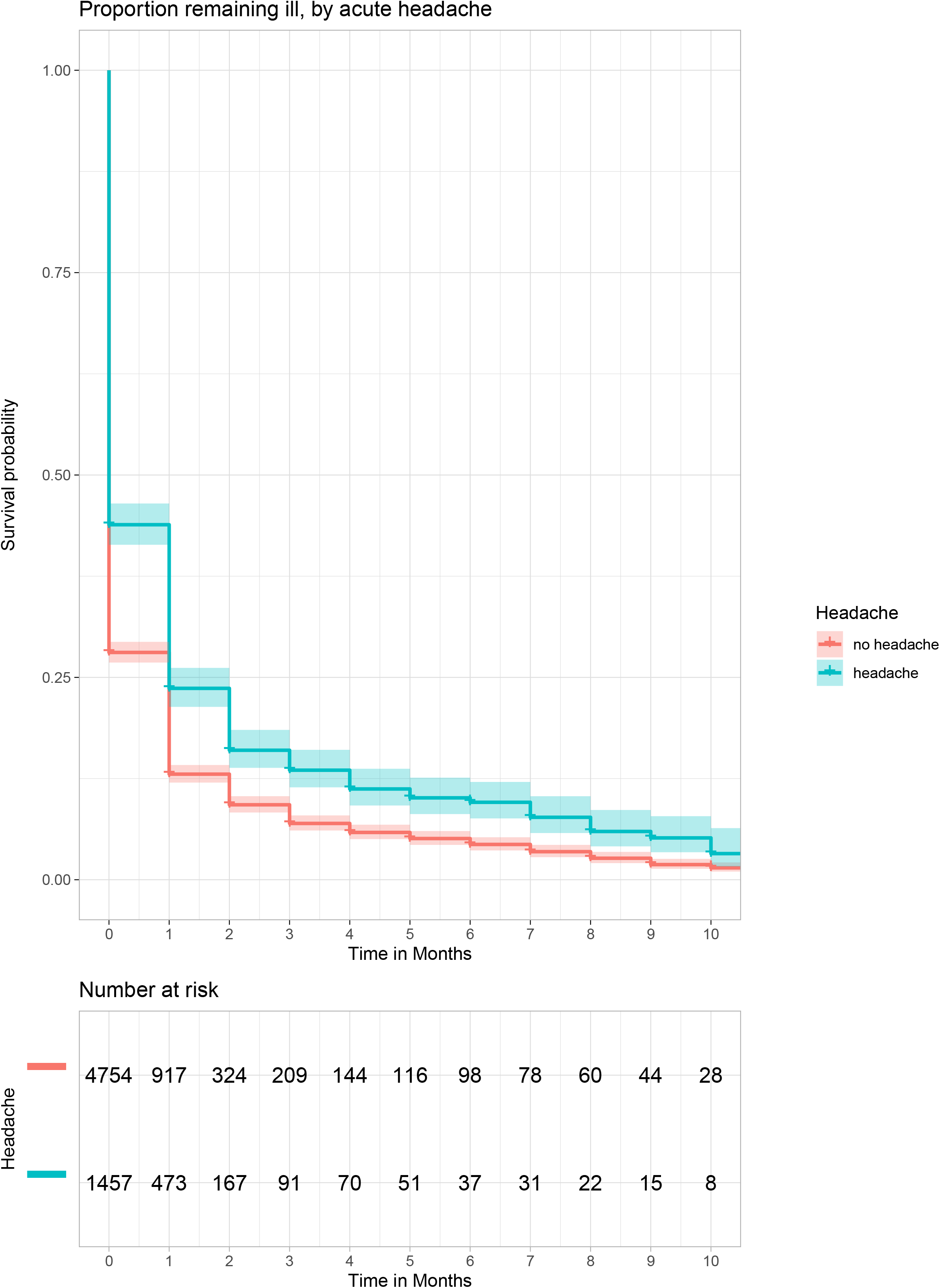
Survival analysis illustrating time to symptom resolution, stratified by presence of absence of headache

In sensitivity analysis, we further restricted the cohort to those with a positive COVID test result (n=4438). In this group, 332 (7.5%) experienced persistence (including 3.3% for at least 4 months and 2.2% for at least 6 months). Repeating contrasts restricting to these groups yielded similar results (see eTable 2 and eFigure 1); in particular, while no significant differences in sociodemographic features were identified, magnitude of association for headache (95% CI 1.43, 95% CI 1.05-1.94) and fever (OR 0.72, 95% CI 0.54-0.95) were similar to that observed in the full cohort.

## DISCUSSION

Across 10 survey waves encompassing 6,211 individuals with COVID-19 illness confirmed by test and/or clinical evaluation, 7.9% experienced persistent symptoms for at least 2 months, and as many as 10 months. Prevalence of persistent symptoms was generally similar across sociodemographic groups, but characteristics of acute illness – in particular, more severe illness and presence of headache −- were associated with greater probability of persistence.

Our results are difficult to compare directly to prior reports. In the largest study to date, at 6-month follow-up, more than half of the cohort experienced persistent symptoms, with fatigue or muscle weakness seen most commonly^4(p19)^. Far less is known about less severely ill individuals. A study using app-based symptom recording in 4182 cases of COVID-19 only 4.5% reported symptoms for more than 8 weeks, broadly similar to our results – as in the inpatient cohorts, fatigue was among the most common symptoms^5^, along with dyspnea and headache. In general, these two studies using very different methodologies and ascertainment methods support the hypothesis that probability of persistent symptoms is likely to correlate with severity of acute illness.

While we identified only a modest magnitude of association with older age, our results may still help to guide risk stratification if confirmed in additional cohorts. Most notably, clinical presentation may point toward those with greater likelihood of experiencing prolonged symptoms, namely greater severity during initial illness, presence of headache, and absence of fever.

Multiple caveats must be considered in interpreting our findings. First, as the study used pre-empaneled respondents in a non-probability design, we cannot reliably calculate response rate; as such, non-response bias cannot be estimated. However, we note that in other domains these non-probability surveys have closely mirrored results with more traditional designs^10^. Furthermore, our cross-sectional design does not allow more precise estimate of symptom persistence, and relies on participant recall in some cases nearly a year after initial illness. Still, this misclassification should bias our results toward smaller estimates of effect, such that any associations we identify may actually represent conservative estimates. For a subset of participants, we also cannot correctly classify their status, as follow-up is insufficient to allow determination of persistence; we report the features of this group, and include survival curves for time to remit, rather than simply excluding them from all analyses. Finally, and most notably, we rely on self-report of symptoms. Here too, this limitation contributes to misclassification, particularly of individuals who may be predisposed to less reliable reporting of symptoms for any reason. As a result, prospective studies will be necessary to confirm our results.

Despite these limitations, we also emphasize the strengths of this systematic assessment, namely that by design it should be more representative than other single-cohort studies because it captures individuals drawn from every state. Moreover, because the survey is not specifically aimed at individuals with COVID-19 or symptom persistence, it may be less biased toward those with a greater interest in long-term symptoms than (for example) symptom tracking applications^5^. We underscore this point: as recruitment materials did not specify COVID-19 or persistence, our results are less likely to reflect individuals with greater interest in COVID-19 persistence.

In aggregate, our results provide further support for the observation that a subset of individuals may experience persistence of COVID-19 symptoms for many months beyond initial infection, providing estimates of prevalence that complement those from hospital-based cohorts or patient surveys. Notably, persistent symptoms are not strongly associated with any individual sociodemographic group; however, individual symptoms and greater overall acuity identify individuals at greater risk for persistence. If confirmed in prospective studies, these results may facilitate risk stratification, with a goal of early intervention to minimize the impact of such Post-acute sequelae of COVID-19, and potentially efforts to prevent them.

## Data Availability

Data are available upon reasonable request.

## Acknowledgements

Dr. Perlis has received consulting fees from Burrage Capital, Genomind, RID Ventures, and Takeda. He holds equity in Outermost Therapeutics and Psy Therapeutics. The other authors report no disclosures.

## Data Sharing Statement

Data are available upon reasonable request.

## Supplemental Materials

eTable 1. Features of individuals who did or did not report COVID-19 symptom persistence, including those who could not be classified

eTable 2. Features of individuals who did or did not report COVID-19 symptom persistence, including those who could not be classified; Limited to participants a self-reported positive COVID-19 test result

eFigure 1. Among those with prior self-reported COVID-19 illness and a positive COVID-19 test, logistic regression model of risk for persistence

## Footnotes

1 The link will be made available after submission.

