## Supplemental Material for "Persistence of symptoms up to 10 months following acute COVID-19 illness"

**eTable 1. Features of individuals who did or did not report COVID-19 symptom persistence, including those who could not be** **classified**

|  | **Resolution in 0-1 month (N=4946)** | **Persistence 2+ months (N=491)** | **Censored (N=774)** | **Total (N=6211)** | **p value** |
| --- | --- | --- | --- | --- | --- |
| **Positive test (a)** | 3424 (69.8%) | 332 (68.3%) | 682 (88.1%) | 4438 (72.0%) | < 0.001 |
| **Clinician diagnosis** | 4565 (92.3%) | 449 (91.4%) | 719 (92.9%) | 5733 (92.3%) | 0.641 |
| **Age (SD)** | 37.81 (11.88) | 38.46 (12.33) | 37.61 (14.07) | 37.83 (12.21) | 0.451 |
| **Gender (Female)** | 2031 (41.1%) | 217 (44.2%) | 553 (71.4%) | 2801 (45.1%) | < 0.001 |
| **Income decile (b)** |  |  |  |  | < 0.001 |
| **0** | 21 (0.4%) | 3 (0.6%) | 8 (1.0%) | 32 (0.5%) |  |
| **1** | 470 (9.5%) | 63 (12.8%) | 84 (10.9%) | 617 (9.9%) |  |
| **2** | 367 (7.4%) | 49 (10.0%) | 97 (12.5%) | 513 (8.3%) |  |
| **3** | 350 (7.1%) | 43 (8.8%) | 92 (11.9%) | 485 (7.8%) |  |
| **4** | 369 (7.5%) | 36 (7.3%) | 91 (11.8%) | 496 (8.0%) |  |
| **5** | 632 (12.8%) | 66 (13.4%) | 128 (16.6%) | 826 (13.3%) |  |
| **6** | 608 (12.3%) | 44 (9.0%) | 110 (14.2%) | 762 (12.3%) |  |
| **7** | 847 (17.2%) | 78 (15.9%) | 80 (10.3%) | 1005 (16.2%) |  |
| **8** | 635 (12.9%) | 59 (12.0%) | 28 (3.6%) | 722 (11.6%) |  |
| **9** | 639 (12.9%) | 50 (10.2%) | 55 (7.1%) | 744 (12.0%) |  |
| **Race** |  |  |  |  | 0.126 |
| **White** | 3697 (74.7%) | 346 (70.5%) | 549 (70.9%) | 4592 (73.9%) |  |
| **Hispanic** | 481 (9.7%) | 46 (9.4%) | 88 (11.4%) | 615 (9.9%) |  |
| **Black** | 470 (9.5%) | 62 (12.6%) | 88 (11.4%) | 620 (10.0%) |  |
| **Asian** | 153 (3.1%) | 17 (3.5%) | 23 (3.0%) | 193 (3.1%) |  |
| **Other** | 145 (2.9%) | 20 (4.1%) | 26 (3.4%) | 191 (3.1%) |  |
| **Residence** |  |  |  |  | < 0.001 |
| **Rural** | 688 (13.9%) | 74 (15.1%) | 134 (17.3%) | 896 (14.4%) |  |
| **Suburban** | 2521 (51.0%) | 270 (55.0%) | 464 (59.9%) | 3255 (52.4%) |  |
| **Urban** | 1737 (35.1%) | 147 (29.9%) | 176 (22.7%) | 2060 (33.2%) |  |
| **Severity (c)** |  |  |  |  | < 0.001 |
| **Not at all** | 724 (14.6%) | 31 (6.3%) | 79 (10.2%) | 834 (13.4%) |  |
| **Not too** | 1337 (27.0%) | 84 (17.2%) | 267 (34.5%) | 1688 (27.2%) |  |
| **Somewhat** | 1673 (33.8%) | 190 (38.9%) | 322 (41.7%) | 2185 (35.2%) |  |
| **Very** | 1210 (24.5%) | 184 (37.6%) | 105 (13.6%) | 1499 (24.2%) |  |
| **Symptoms** |  |  |  |  |  |
| **Fever** | 2434 (49.2%) | 241 (49.1%) | 401 (51.8%) | 3076 (49.5%) | 0.397 |
| **Chills** | 1516 (30.7%) | 209 (42.6%) | 393 (50.8%) | 2118 (34.1%) | < 0.001 |
| **Shaking** | 1392 (28.1%) | 201 (40.9%) | 387 (50.0%) | 1980 (31.9%) | < 0.001 |
| **Congestion** | 2305 (46.6%) | 262 (53.4%) | 483 (62.4%) | 3050 (49.1%) | < 0.001 |
| **Muscle Pain** | 1574 (31.8%) | 223 (45.4%) | 385 (49.7%) | 2182 (35.1%) | < 0.001 |
| **Cough** | 1778 (35.9%) | 199 (40.5%) | 421 (54.4%) | 2398 (38.6%) | < 0.001 |
| **Sore Throat** | 1966 (39.7%) | 226 (46.0%) | 475 (61.4%) | 2667 (42.9%) | < 0.001 |
| **Headache** | 1036 (20.9%) | 167 (34.0%) | 254 (32.8%) | 1457 (23.5%) | < 0.001 |
| **Shortness of Breath** | 1682 (34.0%) | 203 (41.3%) | 365 (47.2%) | 2250 (36.2%) | < 0.001 |
| **Loss of Taste/Smell** | 2437 (49.3%) | 269 (54.8%) | 521 (67.3%) | 3227 (52.0%) | < 0.001 |

**eTable 2. Features of individuals who did or did not report COVID-19 symptom persistence, including those who could not be classified; Limited to participants a self-reported positive COVID-19 test result**

|  | **Resolution in < 2 months (N=3424)** | **Persistence 2+ months (N=332)** | **Ongoing, < 2 months (N=682)** | **Total (N=4438)** | **p value** |
| --- | --- | --- | --- | --- | --- |
| **Positive test (a)** | 3424 (100.0%) | 332 (100.0%) | 682 (100.0%) | 4438 (100.0%) |  |
| **Clinician diagnosis** | 3043 (88.9%) | 290 (87.3%) | 627 (91.9%) | 3960 (89.2%) | 0.032 |
| **Age (SD)** | 36.80 (12.78) | 37.40 (13.16) | 37.39 (14.23) | 36.93 (13.04) | 0.44 |
| **Gender (Female)** | 1757 (51.3%) | 170 (51.2%) | 515 (75.5%) | 2442 (55.0%) | < 0.001 |
| **Income decile (a)** |  |  |  |  | < 0.001 |
| **0** | 18 (0.5%) | 3 (0.9%) | 8 (1.2%) | 29 (0.7%) |  |
| **1** | 357 (10.4%) | 46 (13.9%) | 70 (10.3%) | 473 (10.7%) |  |
| **2** | 312 (9.1%) | 40 (12.0%) | 83 (12.2%) | 435 (9.8%) |  |
| **3** | 304 (8.9%) | 29 (8.7%) | 86 (12.6%) | 419 (9.5%) |  |
| **4** | 317 (9.3%) | 29 (8.7%) | 85 (12.5%) | 431 (9.7%) |  |
| **5** | 508 (14.9%) | 52 (15.7%) | 117 (17.2%) | 677 (15.3%) |  |
| **6** | 442 (12.9%) | 30 (9.0%) | 98 (14.4%) | 570 (12.9%) |  |
| **7** | 506 (14.8%) | 36 (10.8%) | 72 (10.6%) | 614 (13.9%) |  |
| **8** | 320 (9.4%) | 38 (11.4%) | 22 (3.2%) | 380 (8.6%) |  |
| **9** | 334 (9.8%) | 29 (8.7%) | 41 (6.0%) | 404 (9.1%) |  |
| **Race** |  |  |  |  | 0.267 |
| **White** | 2415 (70.5%) | 218 (65.7%) | 491 (72.0%) | 3124 (70.4%) |  |
| **Hispanic** | 421 (12.3%) | 39 (11.7%) | 82 (12.0%) | 542 (12.2%) |  |
| **Black** | 354 (10.3%) | 46 (13.9%) | 71 (10.4%) | 471 (10.6%) |  |
| **Asian** | 114 (3.3%) | 13 (3.9%) | 14 (2.1%) | 141 (3.2%) |  |
| **Other** | 120 (3.5%) | 16 (4.8%) | 24 (3.5%) | 160 (3.6%) |  |
| **Residence** |  |  |  |  | < 0.001 |
| **Rural** | 542 (15.8%) | 54 (16.3%) | 124 (18.2%) | 720 (16.2%) |  |
| **Suburban** | 1829 (53.4%) | 190 (57.2%) | 416 (61.0%) | 2435 (54.9%) |  |
| **Urban** | 1053 (30.8%) | 88 (26.5%) | 142 (20.8%) | 1283 (28.9%) |  |
| **Severity (b)** |  |  |  |  | < 0.001 |
| **Not at all** | 532 (15.5%) | 19 (5.8%) | 66 (9.7%) | 617 (13.9%) |  |
| **Not too** | 1026 (30.0%) | 60 (18.2%) | 250 (36.7%) | 1336 (30.1%) |  |
| **Somewhat** | 1184 (34.6%) | 130 (39.4%) | 282 (41.4%) | 1596 (36.0%) |  |
| **Very** | 680 (19.9%) | 121 (36.7%) | 83 (12.2%) | 884 (19.9%) |  |
| **Symptoms** |  |  |  |  |  |
| **Fever** | 1753 (51.2%) | 180 (54.2%) | 368 (54.0%) | 2301 (51.8%) | 0.28 |
| **Chills** | 1255 (36.7%) | 160 (48.2%) | 357 (52.3%) | 1772 (39.9%) | < 0.001 |
| **Shaking** | 1148 (33.5%) | 149 (44.9%) | 359 (52.6%) | 1656 (37.3%) | < 0.001 |
| **Congestion** | 1786 (52.2%) | 196 (59.0%) | 447 (65.5%) | 2429 (54.7%) | < 0.001 |
| **Muscle Pain** | 1283 (37.5%) | 169 (50.9%) | 354 (51.9%) | 1806 (40.7%) | < 0.001 |
| **Cough** | 1505 (44.0%) | 160 (48.2%) | 391 (57.3%) | 2056 (46.3%) | < 0.001 |
| **Sore Throat** | 1517 (44.3%) | 168 (50.6%) | 438 (64.2%) | 2123 (47.8%) | < 0.001 |
| **Headache** | 836 (24.4%) | 129 (38.9%) | 227 (33.3%) | 1192 (26.9%) | < 0.001 |
| **Shortness of Breath** | 1374 (40.1%) | 155 (46.7%) | 340 (49.9%) | 1869 (42.1%) | < 0.001 |
| **Loss of Taste/Smell** | 1813 (52.9%) | 198 (59.6%) | 480 (70.4%) | 2491 (56.1%) | < 0.001 |
| a. missing 6 (short), 6 (long), 0 (censored) | | |  |  |  |
| b. missing 2 (short), 2 (long), 1 (censored) | | |  |  |  |

**eFigure 1. Among those with prior self-reported COVID-19 illness and a positive COVID-19 test, logistic regression model of risk for persistence**

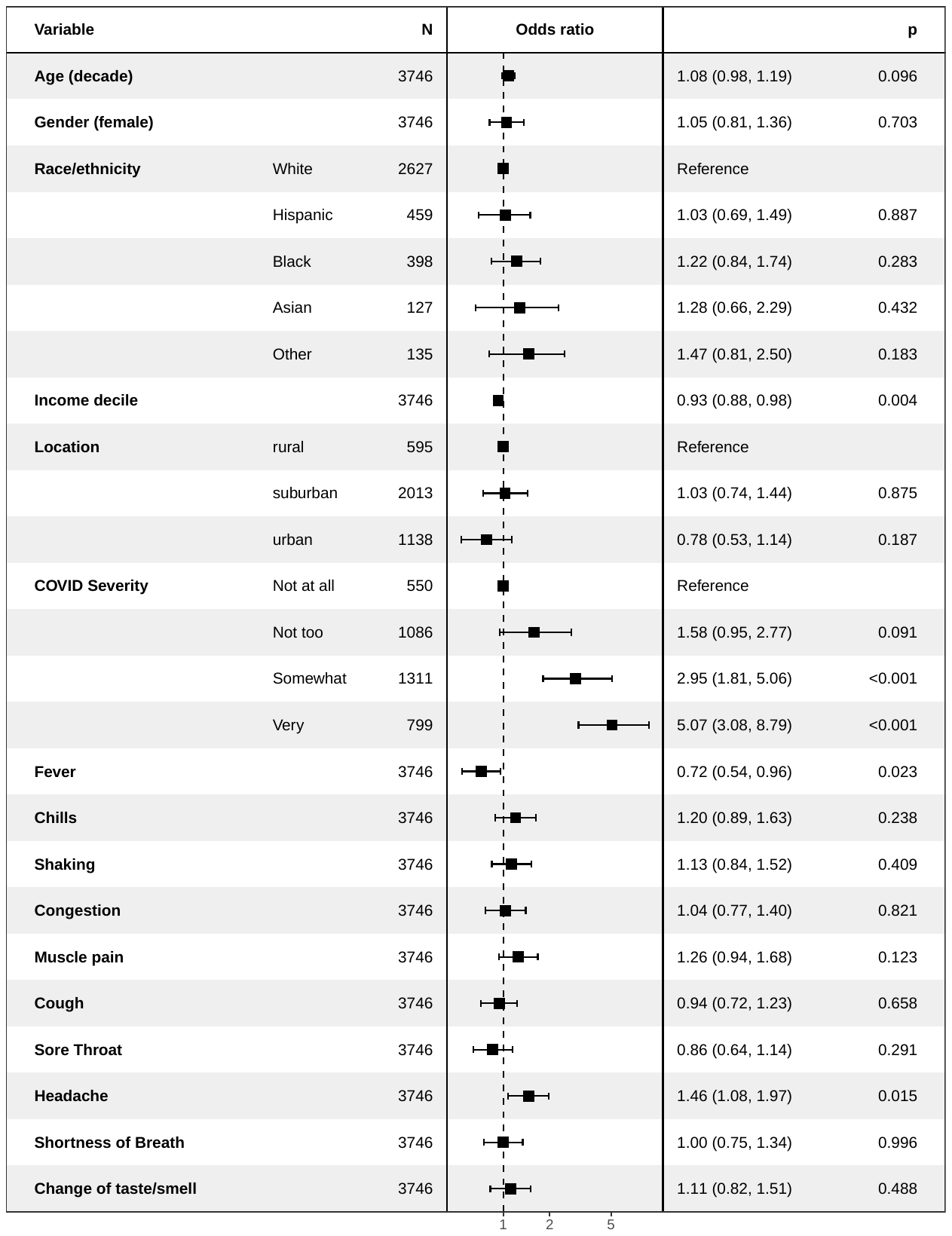
